# A multi-hospital, clinician-initiated bacterial genomics program to investigate treatment failure in severe *Staphylococcus aureus* infections

**DOI:** 10.1101/2023.10.23.23297384

**Authors:** Stefano G. Giulieri, Marcel Leroi, Diane Daniel, Roy Chean, Katherine Bond, Harry Walker, Natasha E. Holmes, Nomvuyo Mothobi, Adrian Alexander, Adam Jenney, Carolyn Beckett, Andrew Mahony, Kerrie Stevens, Norelle L. Sherry, Benjamin P. Howden

**Affiliations:** Department of Microbiology and Immunology, The University of Melbourne at the Doherty Institute for Infection and Immunity, Melbourne, VIC 3000, Australia; Victorian Infectious Disease Service, The Royal Melbourne Hospital, at the Peter Doherty Institute for Infection and Immunity, Melbourne, VIC 3000, Australia; Department of Infectious Diseases, Austin Health, Heidelberg, VIC 3084, Australia; Microbiological Diagnostic Unit Public Health Laboratory, The University of Melbourne at the Doherty Institute for Infection and Immunity, Melbourne, VIC 3000, Australia; Department of Microbiology, Eastern Health, Box Hill, VIC 3128, Australia; Department of Infectious Diseases, Eastern Health, Box Hill, VIC 3128, Australia; Department of Microbiology, Royal Melbourne Hospital, Parkville, VIC 3052, Australia; Department of Microbiology, Western Health, Footscray, VIC 3011, Australia; Department of Infectious Diseases, The University of Melbourne at the Peter Doherty Institute for Infection and Immunity, Melbourne, VIC 3000, Australia; Department of Infectious Diseases, University Hospital Geelong, Geelong, VIC 3220, Australia; Department of Infectious Diseases, The Alfred Hospital, Melbourne, VIC 3004, Australia; Microbiology Unit, The Alfred Hospital, Melbourne, VIC 3004, Australia; Epworth HealthCare, Richmond, VIC 3121, Australia; Department of Infectious Diseases, Bendigo Health, Bendigo, VIC 3550, Australia; Centre for Pathogen Genomics, The University of Melbourne, Melbourne, VIC 3000, Australia

## Abstract

Bacterial genomics is increasingly used for infectious diseases surveillance, outbreak control and prediction of antibiotic resistance. With expanding availability of rapid whole-genome sequencing, bacterial genomics data could become a valuable tool for clinicians managing bacterial infections, driving precision medicine strategies. Here, we present a novel clinician-driven bacterial genomics framework that applies within-patient evolutionary analysis to identify in real-time microbial genetic changes that have an impact on the outcome of severe *Staphylococcus aureus* infections, a strategy that is increasingly used in cancer genomics. Our approach uses a combination of bacterial genomics and novel microbiological testing to identify and track bacterial adaptive mutations that underlie antibiotic treatment failure. We show real-life examples of the impact of our approach and propose a roadmap for the use of bacterial genomics to advance the management of severe bacterial infections.

## Introduction

Thanks to progress in high-throughput whole-genome sequencing (WGS), bacterial genomics has transformed public health microbiology and hospital infection control (Ballard, Sherry, and Howden 2023), and is increasingly used to predict antibiotic resistance (Sherry et al. 2023). However, compared to human and cancer genomics, bacterial genomics has rarely found its application in the clinical setting. This is partly due to turn-around-times that are not suitable for acute infections but also due to still insufficient evidence regarding the impact of bacterial genetic factors on clinical outcomes (Giulieri, Tong, and Williamson 2020).

However, the high degree of resolution achieved through bacterial WGS enables an accurate characterisation of the patient’s individual bacterial strains, which could inform precision infectious diseases management. One of the most promising insights delivered by bacterial genomics is an understanding of the unique evolutionary trajectory of infecting strains during clinical infection. Adaptive evolution plays a key role in bacterial infections, driving antibiotic resistance, infection persistence and immune evasion (Giulieri et al. 2022), yet it is not routinely assessed in clinical practice. This is in contrast to cancer management, where genomics is used to detect mutations acquired *de novo* by cancer cells either at diagnosis or during treatment (Andre et al. 2022).

Here we propose a bacterial genomics framework to assess adaptive evolution during persistent or recurrent infections. We focus on invasive *Staphylococcus aureus* infections, due to the clinical relevance of persistence (Kuehl et al. 2020)(Holland, Bayer, and Fowler 2022) and recurrence (Choi et al. 2021). We hypothesise that bacterial genomics (supported by phenotypic testing) can assist clinicians in determining the cause of treatment failure in *S. aureus* infections, and potentially guide salvage treatments (Holland, Bayer, and Fowler 2022). This conceptual framework is outlined in Figure 1 and Table 1. First, an accurate determination of the genetic distance between isolates collected at baseline and at treatment failure can help distinguish reinfection from relapse, which require different management strategies. Second, genomics and specialised antibiotic susceptibility testing can reveal previously unrecognised resistance mechanisms. Third, meticulous within-host evolution analysis (both phenotypic and molecular) can identify signatures of adaptive evolution, particularly to antibiotics, information that could be useful when selecting a salvage regimen. Finally, if the above investigations remain negative, this suggests lack of adaptive evolution. This finding supports continuing the same antibiotic regimen but suggests the presence of a persistent focus which might warrant more aggressive source control or increase in the antibiotic dose.

**Figure 1.**
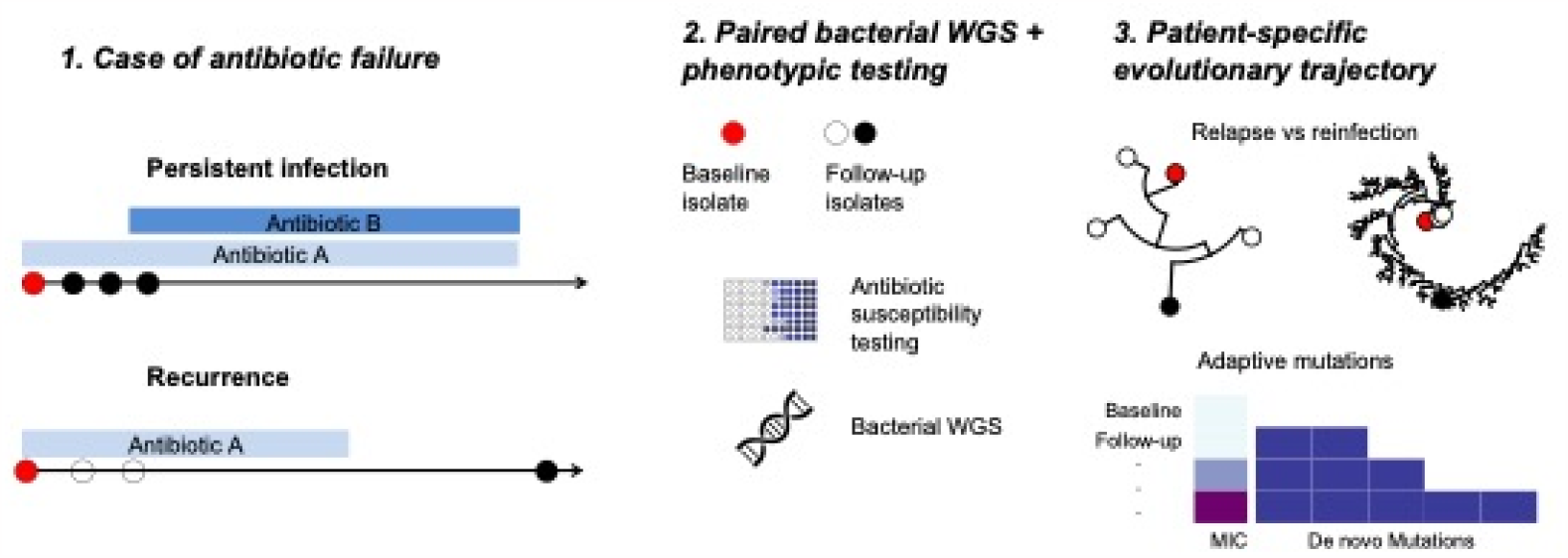
Proposed approach to identify and track bacterial adaptive mutations underlying antibiotic treatment failure.

**Table 1:** Analysis framework.

| ***Clinical question*** | **Bacterial genomics information** | **Potential clinical impact** |
| --- | --- | --- |
| *Reinfection with new strain/clone?* | Large genetic distance between baseline and follow-up strain | Look for source of reinfection – No change of treatment |
| *Antibiotic resistance not detected by routine testing* | Detection of resistance genes/mutations in both baseline and follow-up strain | Change antibiotic treatment |
| *Antibiotic adaptation* | Detection of 'driver mutations' in the follow-up strain | Consider change antibiotic substance or class (consider cross-resistance) |
| *Persistent infection focus?* | Minimal genetic distance between baseline and follow-up strain | Aggressive focus search, source control, increase antibiotic dose |

## Methods

In a preliminary study we offered our genomics investigation framework to teaching hospitals in the state of Victoria, Australia between 2020 and 2023. The investigations were initiated by clinicians in cases of suspected antibiotic treatment failure in invasive *S. aureus* infections indicated by microbiological persistence or recurrence, or unusual phenotypic characteristics of clinical strains noted by medical microbiologists (e.g. increase in minimum inhibitory concentration [MIC], small colony variants). Strains were referred to the Microbiological Diagnostic Unit Public Health Laboratory in Melbourne, where they underwent broth microdilution antibiotic susceptibility testing using the Sensititre® GPN3F and GN6F panels and EUCAST interpretive breakpoints. In addition the cefazolin inoculum effect was assessed as described in (Nannini et al. 2009). Briefly, cefazolin MICs were determined using broth microdilution with a standard (5x10^5^ cfu/ml) and high inoculum (5x10^7^ cfu/ml) dilution. *S. aureus* ATCC 25923 was used as the negative control strain. A > 4-fold increase in the cefazolin MIC with the high inoculum compared to the MIC obtained with the standard inoculum was considered evidence of the cefazolin inoculum effect. Bacterial whole-genome sequencing of same-episode strains was performed as previously described (Giulieri et al. 2022). Single colonies were sequenced using a Illumina Nextseq instrument. Quality control and reads assembly was performed using a standardised pipeline (https://github.com/MDU-PHL/mdu-tools/blob/master/bin/mdu-qc) that calculates reads depth and quality, computes the fraction of *S. aureus* reads using Kraken and assembles reads using Shovill (https://github.com/tseemann/shovill), based on SPAdes (Bankevich et al. 2012). Assemblies were annotated with Prokka (Seemann 2014). Multi-locus sequence type (MLST) was inferred from the assembled contigs using the mlst tool (https://github.com/tseemann/mlst) and resistance genes were detected with Abricate (https://github.com/tseemann/abricate) using the NCBI AMRFinderPlus database (Feldgarden et al. 2021). The within-host evolution genomic analysis (i.e. comparative genomics of bacterial isolates collected at time of failure vs. baseline isolates) was performed using a bespoke pipeline as described in (Giulieri et al. 2022) and available at https://github.com/stefanogg/staph_adaptation_paper. This pipeline uses Snippy (https://github.com/tseemann/snippy) to call variants using the baseline strain draft assembly as a reference. To increase the accuracy of the variant calling, additional filtering steps are added (Giulieri et al. 2018).

Clinicians were provided with two reports: the first included standard antimicrobial susceptibility and the cefazolin inoculum effect results, plus basic genomic characterisation (MLST and resistance gene detection). The second report described the within-host evolutionary analysis and proposed an interpretation based on the structured approach described above (Table 1) and available published evidence regarding the identified mutations.

## Results

From May 2019 to August 2023, we received 65 clinical strains from 13 episodes of invasive *S. aureus* infections from 7 hospitals (median 2 strains per episode, interquartile range 2-4). Details of each episode are provided in Table 2. In all cases, the clinical syndrome was *S. aureus* bacteraemia, of which 9 were recurrent and 2 persistent (in two cases only baseline strains were referred to investigate mechanisms of antibiotic resistance). The investigation was initiated by the treating clinician in 8 cases and by the clinical microbiologist in 5. Antimicrobial susceptibility testing (broth microdilution) showed an increase in oxacillin MIC (≥ 4-fold) between the baseline and subsequent isolates in 2 cases, high baseline oxacillin MIC (1 mg/l) in 3 cases and heteroresistant methicillin-resistant *S. aureus* (MRSA) in one case (oxacillin MIC 0.38/16/32 mg/l). Cefazolin effect testing was done in 3 cases and was found to be present in one.

**Table 2:**
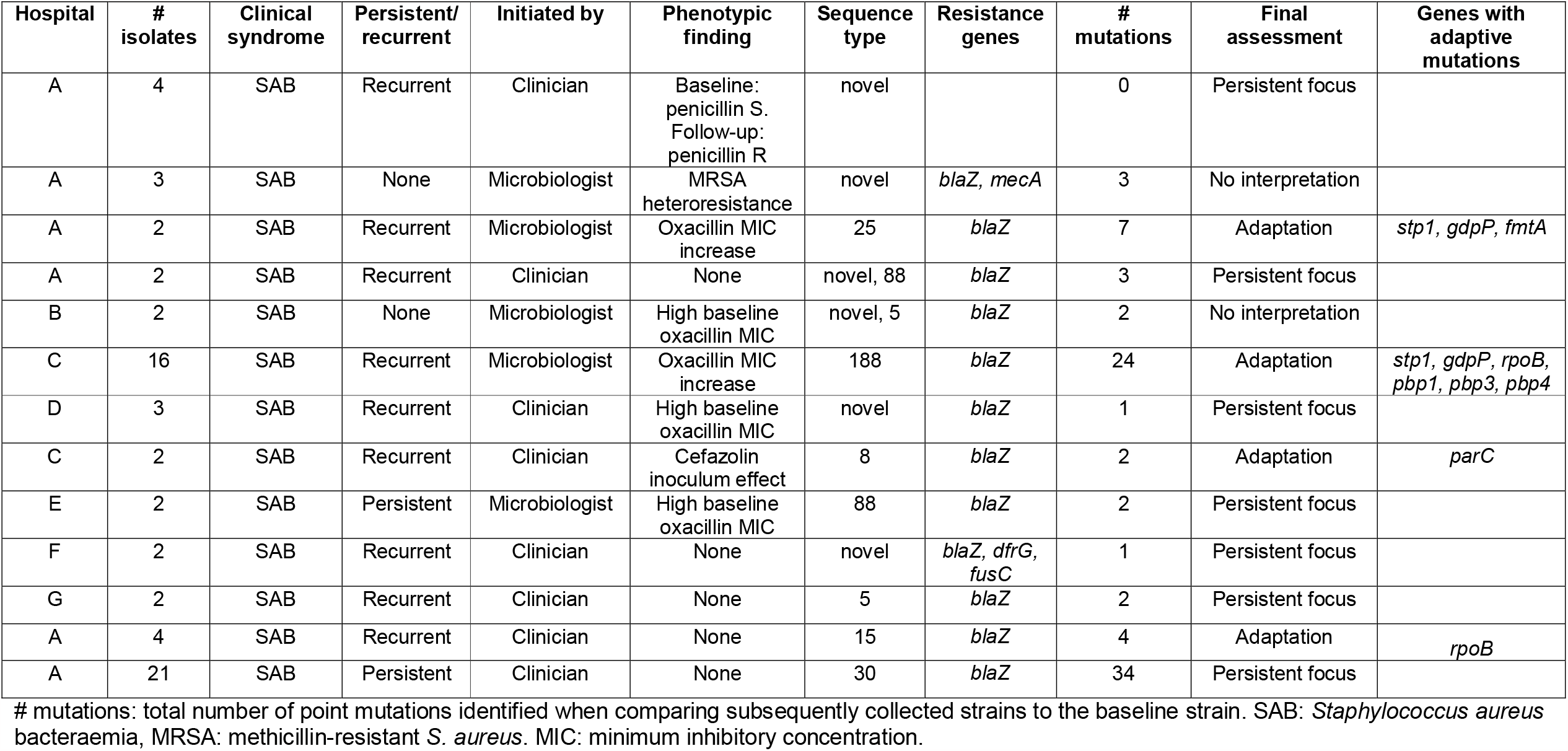
Description of cases.

| Hospital | # isolates | Clinical syndrome | Persistent/recurrent | Initiated by | Phenotypic finding | Sequence type | Resistance genes | # mutations | Final assessment | Genes with adaptive mutations |
| --- | --- | --- | --- | --- | --- | --- | --- | --- | --- | --- |
| A | 4 | SAB | Recurrent | Clinician | Baseline: penicillin S.<br>Follow-up: penicillin R | novel |  | 0 | Persistent focus |  |
| A | 3 | SAB | None | Microbiologist | MRSA heteroresistance | novel | *blaZ*, *mecA* | 3 | No interpretation |  |
| A | 2 | SAB | Recurrent | Microbiologist | Oxacillin MIC increase | 25 | *blaZ* | 7 | Adaptation | *stp1*, *gdpP*, *fmtA* |
| A | 2 | SAB | Recurrent | Clinician | None | novel, 88 | *blaZ* | 3 | Persistent focus |  |
| B | 2 | SAB | None | Microbiologist | High baseline oxacillin MIC | novel, 5 | *blaZ* | 2 | No interpretation |  |
| C | 16 | SAB | Recurrent | Microbiologist | Oxacillin MIC increase | 188 | *blaZ* | 24 | Adaptation | *stp1*, *gdpP*, *rpoB*, *pbp1*, *pbp3*, *pbp4* |
| D | 3 | SAB | Recurrent | Clinician | High baseline oxacillin MIC | novel | *blaZ* | 1 | Persistent focus |  |
| C | 2 | SAB | Recurrent | Clinician | Cefazolin inoculum effect | 8 | *blaZ* | 2 | Adaptation | *parC* |
| E | 2 | SAB | Persistent | Microbiologist | High baseline oxacillin MIC | 88 | *blaZ* | 2 | Persistent focus |  |
| F | 2 | SAB | Recurrent | Clinician | None | novel | *blaZ*, *dfrG*, *fusC* | 1 | Persistent focus |  |
| G | 2 | SAB | Recurrent | Clinician | None | 5 | *blaZ* | 2 | Persistent focus |  |
| A | 4 | SAB | Recurrent | Clinician | None | 15 | *blaZ* | 4 | Adaptation | *rpoB* |
| A | 21 | SAB | Persistent | Clinician | None | 30 | *blaZ* | 34 | Persistent focus |  |

Basic genomic data and within-host evolution analysis are shown in Table 2. Using a structured interpretation, we categorised antibiotic failure as likely due to persistent focus in 7 cases and bacterial adaptation in 4 cases. In two cases our analysis was not conclusive. No cases of co-infection/re-infection were found. Thus, our approach provided bacterial genomic evidence of within-host adaptive evolution in a third of sequentially collected strains. This highlights the clinical relevance of bacterial adaptation during invasive *S. aureus* infections and the interest to trace it using a strategy of sequencing serially collected strains.

Adaptive mutations were observed in genes that have been linked to adaptive antibiotic resistance. For example, mutations in the cyclic-di-AMP phosphodiesterase *gdpP* were found in two of these cases. These were recently shown by multiple groups to drive non-*mec* mediated oxacillin resistance in *S. aureus* (*mec*-independent oxacillin non susceptible *S. aureus*, MIONSA) (Sommer et al. 2021; Argudín et al. 2018; Ba et al. 2019; Giulieri 2023; Giulieri et al. 2020). Mutations in the regulatory serine-threonine phosphatase *stp1* were detected in two isolates. This gene, which has been linked to vancomycin (Cameron et al. 2016) and oxacillin resistance (Chatterjee, Poon, and Chatterjee 2020), displayed one of the strongest statistical signals of adaptive evolution in our large-scale within-host evolution analysis of almost 400 episodes of *S. aureus* infections (Giulieri et al. 2022).

## Discussion

Treatment failure in *S. aureus* invasive infections is challenging for both patients and clinicians. Our clinician-initiated, genomics-informed highlights the potential contribution of within-host evolution analysis to its investigation. We show that an evolutionary genomics framework can provide useful answers for clinical management, for example by identifying mutations in genes that are associated with pathoadaptation or by providing molecular evidence of infection relapse from a persistent focus.

While the work presented here is preliminary and limited to a small number of episodes and to a bacterial species, it provides the proof-of-concept for a structured approach to identify and track bacterial adaptive mutations underlying antibiotic treatment failure using a combination of bacterial genomics and antibiotic susceptibility testing. Full assessment of clinical impacts and utility were not included in the current study, but importantly need to be addressed in future work.

## Data Availability

All data produced in the present study are available upon reasonable request to the authors

## References

Andre, Fabrice, Thomas Filleron, Maud Kamal, Fernanda Mosele, Monica Arnedos, Florence Dalenc, Marie-Paule Sablin, et al. 2022. “Genomics to Select Treatment for Patients with Metastatic Breast Cancer.” Nature 610 (7931): 343–48. 10.1038/s41586-022-05068-3.

Argudín, M. Angeles, S. Roisin, L. Nienhaus, M. Dodémont, R. D. Mendonça, C. Nonhoff, A. Deplano, and O. Denis. 2018. “Genetic Diversity among Staphylococcus Aureus Isolates Showing Oxacillin and/or Cefoxitin Resistance Not Linked to the Presence of Mec Genes.” Antimicrobial Agents and Chemotherapy 62 (7): e00091–18. 10.1128/AAC.00091-18.

Ba, Xiaoliang, Lajos Kalmar, Nazreen F. Hadjirin, Heidrun Kerschner, Petra Apfalter, Fiona J. Morgan, Gavin K. Paterson, et al. 2019. “Truncation of GdpP Mediates β-Lactam Resistance in Clinical Isolates of Staphylococcus Aureus.” The Journal of Antimicrobial Chemotherapy 74 (5): 1182–91. 10.1093/jac/dkz013.

Ballard, Susan A., Norelle L. Sherry, and Benjamin P. Howden. 2023. “Public Health Implementation of Pathogen Genomics: The Role for Accreditation and Application of ISO Standards.” Microbial Genomics 9 (8): 001097. 10.1099/mgen.0.001097.

Bankevich, Anton, Sergey Nurk, Dmitry Antipov, Alexey A. Gurevich, Mikhail Dvorkin, Alexander S. Kulikov, Valery M. Lesin, et al. 2012. “SPAdes: A New Genome Assembly Algorithm and Its Applications to Single-Cell Sequencing.” Journal of Computational Biology 19 (5): 455–77. 10.1089/cmb.2012.0021.

Cameron, David R., Jhih-Hang Jiang, Xenia Kostoulias, Daniel J. Foxwell, and Anton Y. Peleg. 2016. “Vancomycin Susceptibility in Methicillin-Resistant Staphylococcus Aureus Is Mediated by YycHI Activation of the WalRK Essential Two-Component Regulatory System.” Scientific Reports 6 (1): 30823. 10.1038/srep30823.

Chatterjee, Aditi, Raymond Poon, and Som S. Chatterjee. 2020. “Stp1 Loss of Function Promotes β-Lactam Resistance in Staphylococcus Aureus That Is Independent of Classical Genes.” Antimicrobial Agents and Chemotherapy 64 (6): 10.1128/aac.02222-19. 10.1128/aac.02222-19.

Choi, Seong-Ho, Michael Dagher, Felicia Ruffin, Lawrence P Park, Batu K Sharma-Kuinkel, Maria Souli, Alison M Morse, et al. 2021. “Risk Factors for Recurrent Staphylococcus Aureus Bacteremia.” Clinical Infectious Diseases 72 (11): 1891–99. 10.1093/cid/ciaa801.

Feldgarden, Michael, Vyacheslav Brover, Narjol Gonzalez-Escalona, Jonathan G. Frye, Julie Haendiges, Daniel H. Haft, Maria Hoffmann, et al. 2021. “AMRFinderPlus and the Reference Gene Catalog Facilitate Examination of the Genomic Links among Antimicrobial Resistance, Stress Response, and Virulence.” Scientific Reports 11 (1): 12728. 10.1038/s41598-021-91456-0.

Giulieri, Stefano G. 2023. “Case Commentary: The Hidden Side of Oxacillin Resistance in Staphylococcus Aureus.” Antimicrobial Agents and Chemotherapy 0 (0): e00716–23. 10.1128/aac.00716-23.

Giulieri, Stefano G., Sarah L. Baines, Romain Guerillot, Torsten Seemann, Anders Gonçalves da Silva, Mark Schultz, Ruth C. Massey, Natasha E. Holmes, Timothy P. Stinear, and Benjamin P. Howden. 2018. “Genomic Exploration of Sequential Clinical Isolates Reveals a Distinctive Molecular Signature of Persistent Staphylococcus Aureus Bacteraemia.” Genome Medicine 10 (1): 65. 10.1186/s13073-018-0574-x.

Giulieri, Stefano G, Romain Guérillot, Sebastian Duchene, Abderrahman Hachani, Diane Daniel, Torsten Seemann, Joshua S Davis, et al. 2022. “Niche-Specific Genome Degradation and Convergent Evolution Shaping Staphylococcus Aureus Adaptation during Severe Infections.” Edited by Bavesh D Kana, Daria Van Tyne, and Meiqin Zheng. eLife 11 (June): e77195. 10.7554/eLife.77195.

Giulieri, Stefano G., Romain Guérillot, Jason C. Kwong, Ian R. Monk, Ashleigh S. Hayes, Diane Daniel, Sarah Baines, et al. 2020. “Comprehensive Genomic Investigation of Adaptive Mutations Driving the Low-Level Oxacillin Resistance Phenotype in Staphylococcus Aureus.” Edited by Melinda M. Pettigrew. mBio 11 (6): e02882–20. 10.1128/mBio.02882-20.

Giulieri, Stefano G., Steven Y. C. Tong, and Deborah A. Williamson. 2020. “Using Genomics to Understand Meticillin- and Vancomycin-Resistant Staphylococcus Aureus Infections.” Microbial Genomics. Microbiology Society. 10.1099/mgen.0.000324.

Holland, Thomas L, Arnold S Bayer, and Vance G Fowler. 2022. “Persistent MRSA Bacteremia: Resetting the Clock for Optimal Management.” Clinical Infectious Diseases, May, ciac364. 10.1093/cid/ciac364.

Kuehl, Richard, Laura Morata, Christian Boeing, Isaac Subirana, Harald Seifert, Siegbert Rieg, Winfried V Kern, et al. 2020. “Defining Persistent Staphylococcus Aureus Bacteraemia: Secondary Analysis of a Prospective Cohort Study.” The Lancet Infectious Diseases 20 (12): 1409–17. 10.1016/S1473-3099(20)30447-3.

Nannini, Esteban C., Martin E. Stryjewski, Kavindra V. Singh, Agathe Bourgogne, Tom H. Rude, G. Ralph Corey, Vance G. Fowler, and Barbara E. Murray. 2009. “Inoculum Effect with Cefazolin among Clinical Isolates of Methicillin-Susceptible Staphylococcus Aureusc: Frequency and Possible Cause of Cefazolin Treatment Failure.” Antimicrobial Agents and Chemotherapy 53 (8): 3437–41. 10.1128/AAC.00317-09.

Seemann, Torsten. 2014. “Prokka: Rapid Prokaryotic Genome Annotation.” Bioinformatics 30 (14): 2068–69. 10.1093/bioinformatics/btu153.

Sherry, Norelle L., Kristy A. Horan, Susan A. Ballard, Anders Goncalves da Silva, Claire L. Gorrie, Mark B. Schultz, Kerrie Stevens, et al. 2023. “An ISO-Certified Genomics Workflow for Identification and Surveillance of Antimicrobial Resistance.” Nature Communications 14 (1): 60. 10.1038/s41467-022-35713-4.

Sommer, Anna, Stephan Fuchs, Franziska Layer, Christoph Schaudinn, Robert E. Weber, Hugues Richard, Mareike B. Erdmann, et al. 2021. “Mutations in the gdpP Gene Are a Clinically Relevant Mechanism for β-Lactam Resistance in Meticillin-Resistant Staphylococcus Aureus Lacking Mec Determinants.” Microbial Genomics 7 (9): 000623. 10.1099/mgen.0.000623.

